# Impact of Socioeconomic Status on Clinical Features and Outcomes of Bacterial Keratitis: The Midlands Infectious Keratitis Study

**DOI:** 10.64898/2026.04.07.26350291

**Authors:** Khawaja Muhammad Ammar Ali Javed, Burak Ozturk, Saba Anwar, Gibran Butt, Liying Low, Dalia G. Said, Harminder S Dua, Saaeha Rauz, Darren S. J. Ting

## Abstract

**Background/Aims:** To evaluate the impact of socioeconomic deprivation on clinical presentation and outcomes of bacterial keratitis (BK) in the United Kingdom.

**Methods:** A retrospective multicentre cohort study of 320 patients with BK presenting to two UK tertiary ophthalmic centres. Demographic, clinical and microbiological data were extracted from electronic health records. Socioeconomic status was assigned using residential postcodes mapped to the 2019 English Index of Multiple Deprivation (IMD) and grouped into quintiles (Q1 most deprived; Q5 least deprived). Presenting severity and outcomes were compared across IMD quintiles.

**Results:** The mean age was 54.0±20.9 years; 50.6% were male and 83.4% were White. Mean presenting CDVA was 1.10±1.01 logMAR and time to presentation was a median of 3 days (IQR 1–6). Most cases had a small infiltrate (<3Lmm; 68.4%), small epithelial defect (<3Lmm; 63.4%) and no hypopyon (72.5%). Hospitalisation was required in 50.0%, and 17.5% underwent surgery. Culture positivity was 36.3%. There were no significant differences in presenting CDVA, time to presentation, clinical severity, admission, microbiological profile, surgical intervention or final CDVA across IMD quintiles (all p>0.05). Final CDVA improved to 0.75±0.96 logMAR (p<0.001). On multivariable analysis, poorer final CDVA was associated with worse presenting CDVA, increasing age and Gram-positive organisms, but not IMD.

**Conclusion:** Socioeconomic deprivation did not influence the clinical presentation or outcomes in BK. Clinical severity at presentation and microbiological profile were the principal determinants of outcome. In this acute, painful sight-threatening condition, deprivation-related disparities may be attenuated by prompt presentation and universal access to emergency ophthalmic care.

## INTRODUCTION

Infectious keratitis (IK) is a leading cause of corneal blindness worldwide and an ophthalmic emergency requiring prompt diagnosis and intensive antimicrobial therapy.^1–3^ Despite advances in diagnostics and treatment, IK continues to cause substantial visual morbidity and economic burden.^4,5^ In the United Kingdom (UK), the incidence of IK ranges from 34 to 52 cases per 100,000 per year, with the majority (∼90%) due to bacterial keratitis (BK), commonly associated with contact lens wear and ocular surface disease.^6–8^

A growing body of literature suggests that social determinants of health influence both disease risk and outcomes.^9–11^ Individuals living in deprived areas may face barriers to early medical consultation, reduced access to primary care, and delays in referral to specialist services.^12–14^ Although socioeconomic deprivation has been linked to inequalities in eye-care access and outcomes in the UK, its specific impact on BK remains poorly understood.^11^

In chronic ophthalmic diseases, deprivation-linked disparities have been consistently demonstrated. Studies in glaucoma show that patients from more deprived areas present with significantly worse visual-field loss at diagnosis.^15,16^ Similar trends have been reported in diabetic retinopathy and age-related macular degeneration, where deprivation is associated with later presentation and reduced adherence to follow-up.^17,18^ However, acute ophthalmic infections such as BK differ fundamentally from chronic diseases; they progress rapidly, often within hours to days, and their outcomes depend heavily on time-to-presentation and immediate access to care.^4,19,20^

Recent US data demonstrate that neighbourhood-level deprivation and social risk factors are associated with worse presenting visual acuity in IK, suggesting delayed access to care.^21^ However, comparable UK-based evidence remains limited.^22,23^

The English Index of Multiple Deprivation (IMD) 2019 is a validated small-area measure of socioeconomic status, incorporating domains such as income, employment, health, education, housing, and environment.^24^ It has been widely applied in ophthalmic health-services research, including cataract, glaucoma, and eye-care utilisation studies.^15,16,25–27^

This multicentre study aimed to examine the impact of socioeconomic deprivation, measured using IMD, on presenting clinical severity and outcomes of BK in the UK. Specifically, we aimed to: (1) describe baseline characteristics across IMD quintiles; (2) compare clinical severity, hospitalisation, and outcomes by deprivation level; and (3) assess associations between deprivation and presenting or final vision.

## METHODS

### Study design and population

This retrospective multicentre cohort study included patients with BK presenting to the Birmingham and Midland Eye Centre (BMEC), Birmingham, and Queen’s Medical Centre (QMC), Nottingham, UK, between 2015 and 2020. The study was approved by the clinical governance team of both institutions as a clinical audit (BMEC Ref: 2444; QMC Ref: 19–265C) and was conducted in accordance with the tenets of Declaration of Helsinki. Ethics approval was not required as this clinical audit only involved retrospective study of anonymised data that were collected during the process of standard clinical care.

Consecutive cases were identified from electronic records and de-identified prior to analysis.^23,28^ BK was defined as a corneal epithelial defect with underlying stromal infiltrate and associated inflammation, with or without positive microbiology. Patients required a valid residential postcode for socioeconomic linkage. Cases with alternative diagnoses (e.g. immune-mediated keratitis, sterile corneal infiltrates or ulceration) were excluded.

### Clinical assessment

All patients underwent slit-lamp examination at presentation. Clinical assessment included documentation of infiltrate size, epithelial defect size, stromal involvement, and presence of hypopyon, following standardised UK corneal infection pathways and international recommendations.

### Microbiological investigations

Microbiological sampling was performed for all moderate-to-severe ulcers and for milder cases at clinician discretion, in accordance with departmental guidelines for IK. In line with this protocol, all ulcers >1 mm in diameter, centrally located, associated with significant anterior chamber reaction or hypopyon, or demonstrating atypical features underwent corneal scraping for microscopy and culture. Corneal scrapings were taken using sterile hypodermic needles / swabs and directly inoculated onto chocolate agar (for fastidious organisms), blood agar (for bacterial pathogens), and Sabouraud dextrose agar (for fungi). For suspected cases of *Acanthamoeba* keratitis, a corneal swab and/or epithelial biopsy was obtained for PCR and/or culture on non-nutrient agar with *Escherichia coli* overlay. All cultures were incubated for at least 1 week (and up to 3 weeks for suspected *Acanthamoeba* keratitis and fungal keratitis). These procedures were conducted according to contemporary UK investigative protocols used in previous studies.^4,8,28^

### Treatment protocol

Initial empirical treatment followed local guidelines consistent with national recommendations. All patients with presumed BK were started on intensive hourly topical antimicrobial therapy, typically levofloxacin 0.5% monotherapy for less severe ulcers, or combined fortified therapy using cefuroxime 5% and an aminoglycoside (amikacin 2.5% or gentamicin 1.5%) for moderate-to-severe disease, depending on clinician judgement and ulcer severity. Hospitalisation was warranted for severe ulcers, defined as central involvement, infiltrate >2 mm, presence of hypopyon or when patients were unable or unlikely to comply with the intensive treatment regimen, or when the ulcer failed to respond to initial therapy. All admitted patients were commenced on combined fortified therapy.

Antimicrobial regimens were subsequently adjusted according to microbiological results, antibiotic susceptibility, and clinical response. Topical cycloplegic drops and oral analgesia were routinely administered. Surgical interventions, including corneal gluing, amniotic membrane transplantation, or therapeutic penetrating keratoplasty, were performed when required for non-resolving infection, progressive stromal thinning, or frank corneal perforation.

### Socioeconomic linkage

Residential postcodes were mapped to the English IMD 2019, which were representative of the socioeconomic status for the study time period, using the Office for National Statistics Open Data Communities platform.^24^ IMD deciles were collapsed into quintiles to facilitate statistical interpretation, with quintile 1 representing the most deprived areas and quintile 5 the least deprived.

### Clinical variables and outcomes

Extracted variables included age, sex, race, presenting CDVA (logMAR), time to presentation, infiltrate size, epithelial defect size, hypopyon, microbiological results, organism group, hospital admission, length of stay, and surgical intervention.

Infiltrate and epithelial defect sizes were categorised as small (≤3 mm), moderate (3.1–6 mm), or large (>6 mm).^7^ Predisposing risk factors were classified as contact lens wear, ocular trauma, ocular surface disease, or other (e.g. corticosteroid use, prior surgery, systemic immunosuppression). As multiple risk factors could coexist, each was analysed as a binary variable. Primary outcomes were presenting severity and final CDVA. Secondary outcomes included hospitalisation, length of stay (LOS), and surgical intervention.

### Statistical analysis

Analyses were performed using R (v4.3.0) and Python (v3.13). Continuous variables were summarised as mean ± standard deviation (SD) and median (IQR), and categorical variables as frequencies (%). Presenting and final CDVA were analysed as paired variables using the Wilcoxon signed-rank test due to non-normal distribution (Shapiro–Wilk p<0.0001). Associations between presenting and final CDVA were assessed using Spearman correlation. Comparisons across IMD quintiles used Kruskal–Wallis tests for continuous variables and chi-square or Fisher’s exact tests for categorical variables. Pairwise comparisons between the most and least deprived groups (Q1 vs Q5) used Mann–Whitney U tests or chi-square/Fisher’s exact tests as appropriate.

Multivariable linear and logistic regression models were used to assess associations between deprivation and outcomes, adjusting for age, sex, race, IMD quintile, presenting CDVA (where appropriate), and relevant clinical variables. Statistical significance was defined as p<0.05.

## RESULTS

### Baseline characteristics

A total of 320 eyes from 320 patients were included. Baseline demographic, clinical and microbiological characteristics are presented in **Table 1**. The mean age was 54.0 ± 20.9 years (median 54.5 years; IQR 36.1–71.8), and sex distribution was balanced (49.4% female). Most patients were White (83.4%), with smaller proportions of Asian (7.8%), Black (1.6%), and Other/Unknown race (7.2%).

**Table 1.**
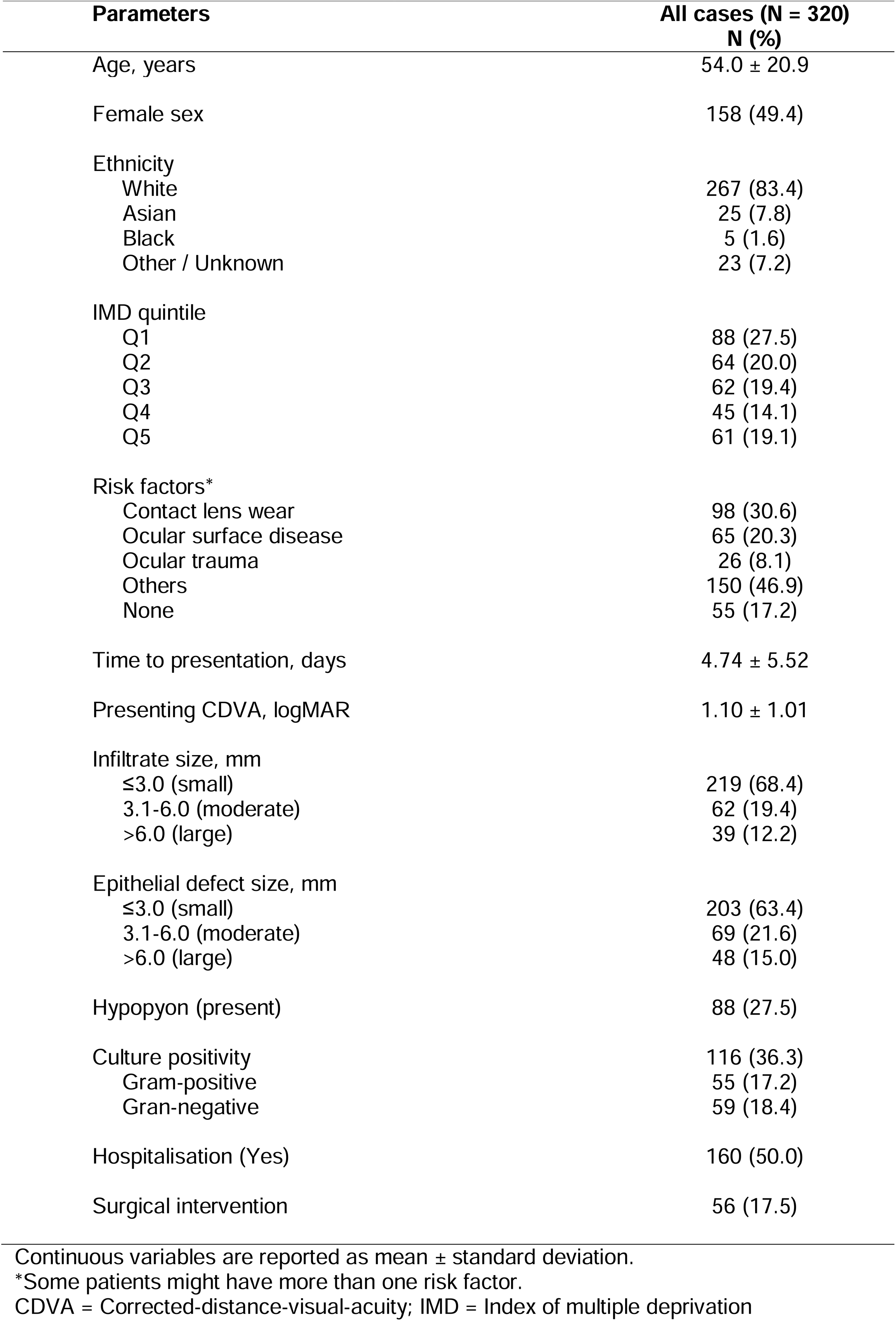
Baseline characteristics of patients with bacterial keratitis.

At presentation, the mean CDVA was 1.10 ± 1.01 logMAR (median 0.70 logMAR; IQR 0.20–1.90); The median time to presentation was 3.0 days (IQR 1.0–6.0). 12.2% of ulcers had a large infiltrate and 15.0% had a large epithelial defect. Hypopyon was present in 27.5% of cases. Half of the cohort (160, 50%) required hospitalisation. Predisposing risk factors were common and not mutually exclusive: contact lens wear was present in 30.6% of cases, ocular trauma in 8.1%, ocular surface disease in 20.3%, and other risk factors in 46.9%, while 17.2% had no identifiable risk factor. Culture positivity was 36.3%, with Gram-negative (18.4%) and Gram-positive organisms (17.2%) isolated at similar frequencies.

### Comparison by socioeconomic status

When patients were stratified by socioeconomic status, there were no significant differences in age, presenting CDVA, time to presentation, final CDVA, or duration of hospitalisation across IMD quintiles (all p>0.05; **Table 2**). The distributions of presenting CDVA and final CDVA by IMD quintile are illustrated in **Figure 1A** and **Figure 1B**, respectively.

**Figure 1.**
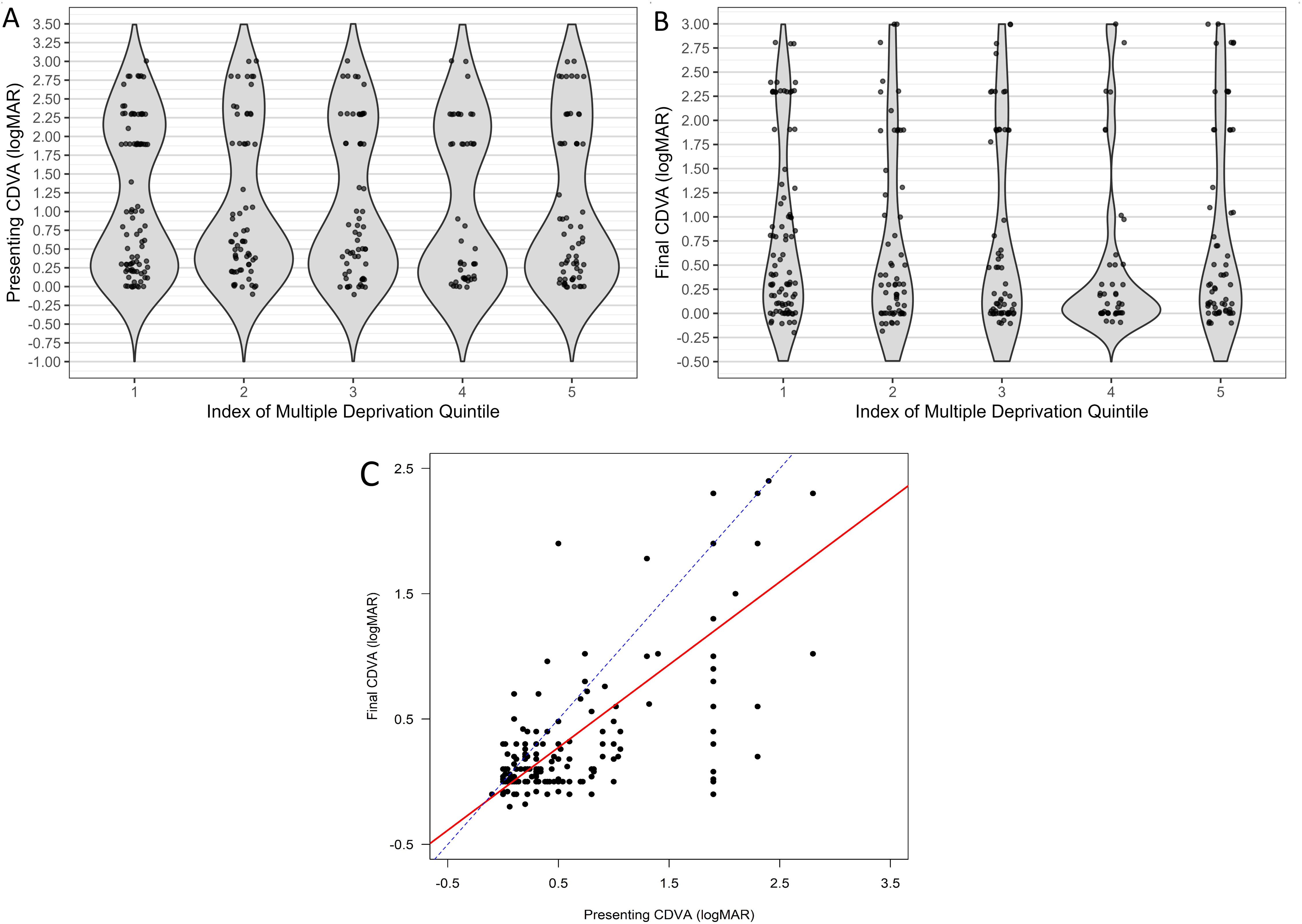
Visual acuity distribution and outcomes in bacterial keratitis across socioeconomic deprivation. (A) Distribution of presenting corrected distance visual acuity (CDVA, logMAR) across Index of Multiple Deprivation (IMD) quintiles (Q1 = most deprived; Q5 = least deprived). Violin plots show data distribution with jittered individual observations. No significant differences were observed between IMD groups (p > 0.05). (B) Distribution of final CDVA (logMAR) across IMD quintiles at discharge or final follow-up, displayed using violin plots with individual data points. Final visual outcomes did not differ significantly across IMD groups (p > 0.05). (C) Relationship between presenting and final CDVA (logMAR). Scatter plot of final CDVA versus presenting CDVA. The red solid line represents the line of best fit, demonstrating a positive association between presenting and final CDVA, while the dashed blue line represents the line of equality (y = x). Points below the line of equality indicate visual improvement, whereas points above indicate deterioration. Poorer presenting CDVA was associated with worse final visual outcomes. CDVA = corrected distance visual acuity; IMD = Index of Multiple Deprivation.

**Table 2.**
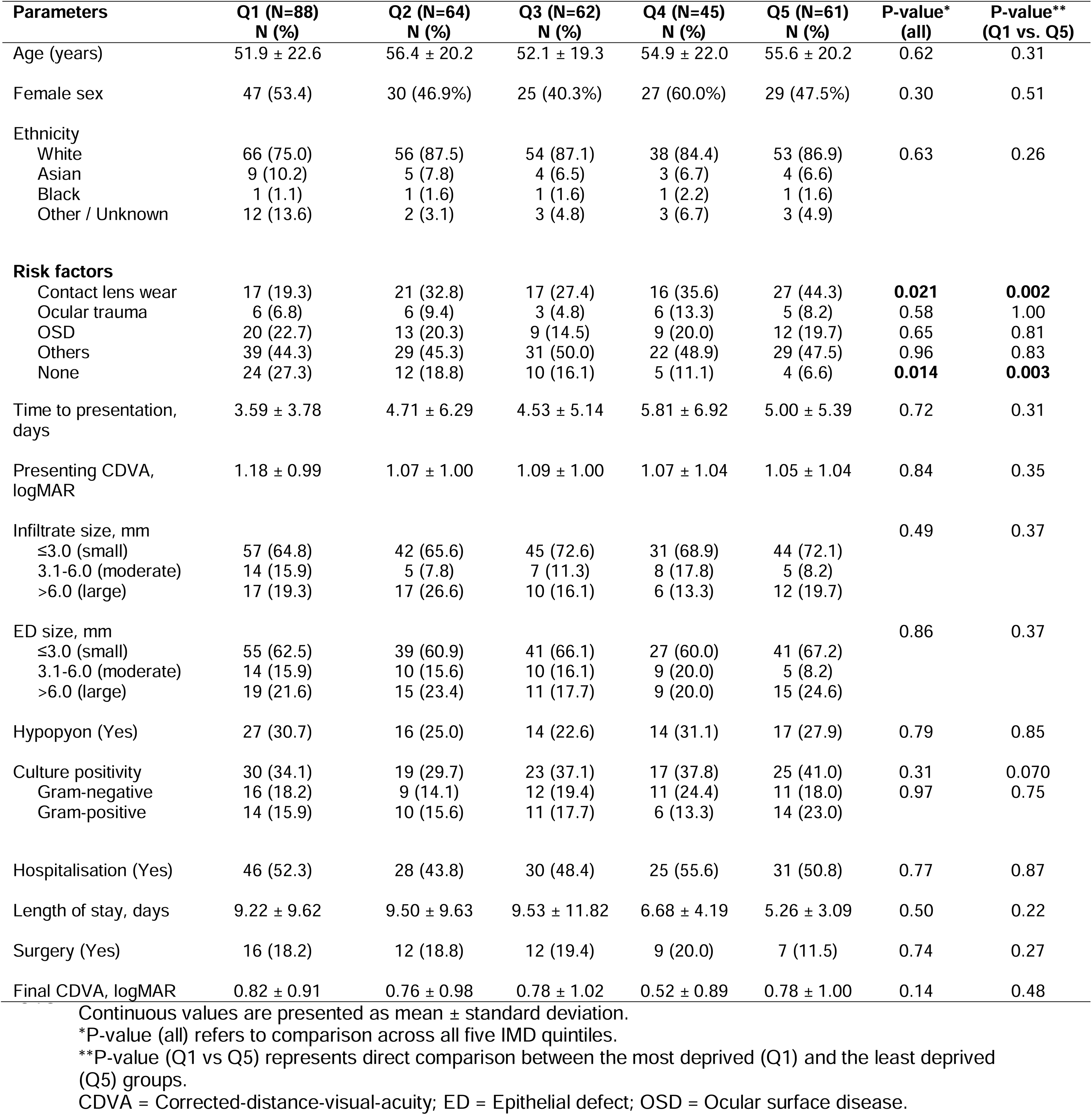
Clinical characteristics stratified by Index of Multiple Deprivation (IMD) quintile.

The distributions of race (p=0.63), infiltrate size (p=0.49), epithelial defect size (p=0.86), hypopyon (p=0.79), culture positivity (p=0.31), organism distribution (p=0.97), admission (p=0.77), and surgical intervention (p=0.74) did not differ significantly across IMD quintiles

(**Table 2**). Risk factors were broadly similar across IMD quintiles, although contact lens wear was significantly more common among less deprived groups (overall p=0.021; Q1 vs. Q5 p=0.002). In univariable comparisons, none of the clinical severity parameters demonstrated a pattern suggesting more advanced disease at presentation in the most deprived group (Q1) relative to the least deprived (Q5) (all p>0.05).

### Potential factors influencing presenting clinical features

Multivariable analyses assessing presenting characteristics are shown in **Table 3**, with full regression outputs provided in **Supplementary Tables S1–S5**. After adjusting for age, sex, race, and IMD quintile, deprivation was not associated with presenting CDVA, with all IMD quintiles showing non-significant effects (e.g., IMD Q1 vs Q5 β 0.243; 95% CI –0.095–0.580; p=0.16). Older age was also associated with poorer presenting CDVA (β=0.017; 95% CI, 0.011–0.023; p<0.001) (**Supplementary Table S1**). When comparing between IMD Q1 and Q5, deprivation was not associated with hypopyon (OR=1.14; 95% CI, 0.52-2.49; p=0.74), large infiltrate (OR=2.08; 95% CI, 0.59-7.30; p=0.25), large epithelial defect (OR=2.03; 95% CI, 0.59–6.94; p=0.26), or hospitalisation (OR=1.14; 95% CI, 0.56-2.31; **Table 3**).

**Table 3.**
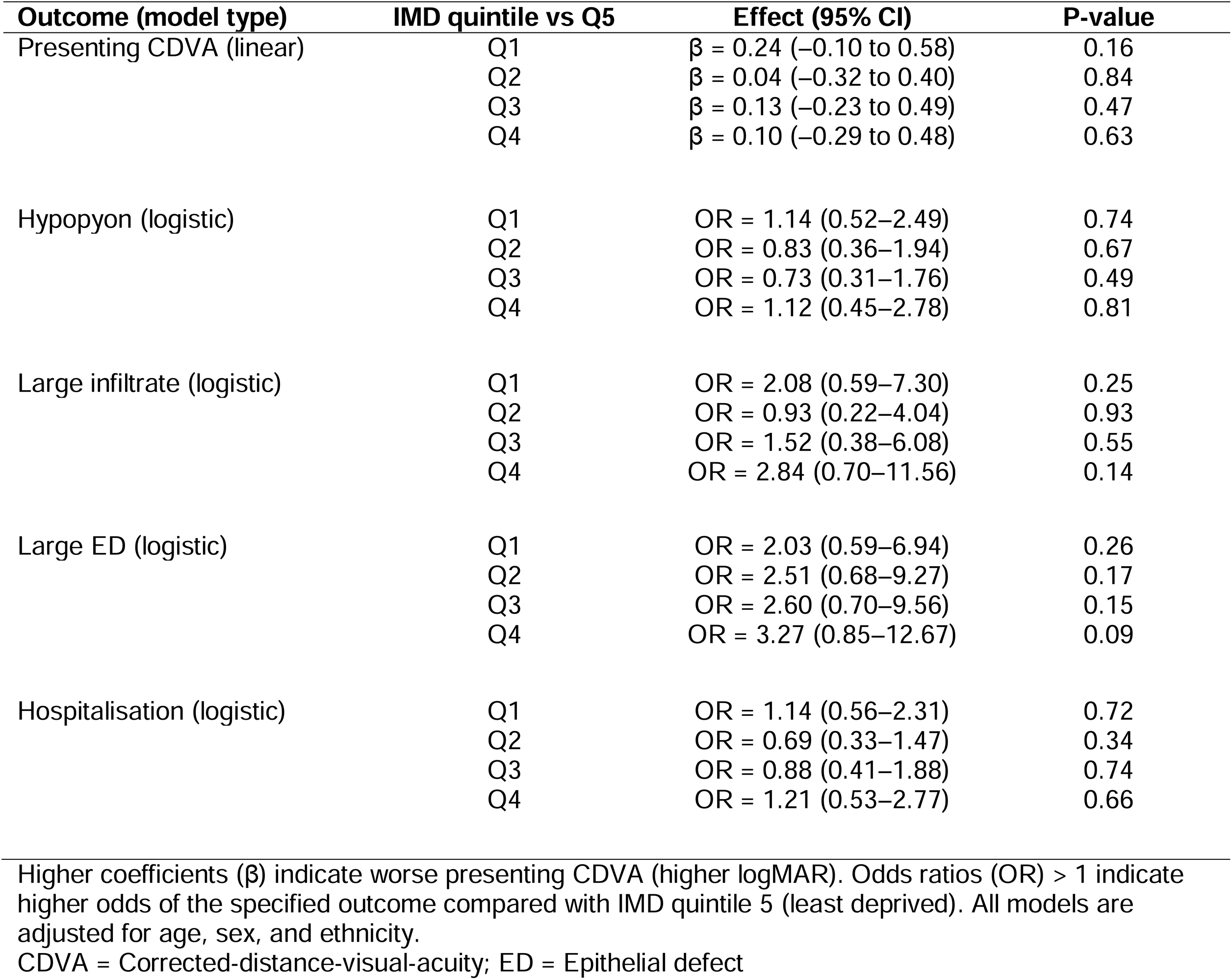
Adjusted associations between Index of Multiple Deprivation (IMD) quintile and presenting clinical severity.

Across all three inflammatory severity models (**Supplementary Table S2-S4**), presenting CDVA was the strongest predictor of clinical severity: poorer presenting vision was associated with substantially higher odds of hypopyon (OR=1.79; 95% CI ,1.37–2.36; p<0.001), large infiltrate (OR =6.10; 95% CI, 3.28–11.32; p<0.001), and large epithelial defect (OR=5.00; 95% CI 3.07–8.15; p<0.001). Admission was not associated with deprivation, however, presenting CDVA remained a strong independent determinant of admission (OR 1.69; 95% CI, 1.30-2.18; p<0.001) (**Supplementary Table S5**).

### Potential factors influencing clinical outcomes

Outcome analyses by IMD quantiles are presented in **Table 4**, with full models shown in **Supplementary Tables S6–8**. Final CDVA improved significantly from presentation (Wilcoxon signed-rank test, p<0.001), decreasing from a mean presenting CDVA of 1.10±1.01 logMAR to a final BCVA of 0.75±0.96 logMAR. Presenting and final CDVA were strongly correlated (Spearman ρ=0.72, p<0.001), indicating that poorer presenting vision was associated with poorer final outcomes. The association between presenting and final CDVA is shown in **Figure 1C**, demonstrating a strong monotonic relationship. IMD quintile was not associated with final CDVA, except for IMD Q4, where patients had slightly better final vision than those in Q5 (β=–0.284; 95% CI, –0.507 to –0.061; p=0.013) (**Supplementary Table S6**). Final CDVA was strongly associated with presenting CDVA and age, indicating that initial visual impairment and older age remained major determinants of visual prognosis. Presenting CDVA showed the largest independent effect on final vision (β=0.628; 95% CI, 0.545–0.710; p<0.001), and older age was similarly associated with worse final CDVA (β 0.008; 95% CI, 0.005–0.012; p<0.001). Microbiological findings were also strongly associated with outcome: Gram-positive organisms were associated with significantly worse final CDVA (β 0.299; 95% CI, 0.076–0.521; p=0.009). In the surgical intervention model (**Supplementary Table S7**), deprivation was not associated with the need for surgery. Instead, culture positivity, particularly Gram-positive was strongly associated with higher surgical risk, as was the presence of a large epithelial defect. For example, Gram-positive organisms increased the odds of surgery nearly fourfold (OR 3.98; 95% CI, 1.19–13.29; p=0.025). Worse presenting CDVA further increased the odds of undergoing surgery (OR 1.83; 95% CI, 1.17–2.85; p=0.008).

**Table 4.**
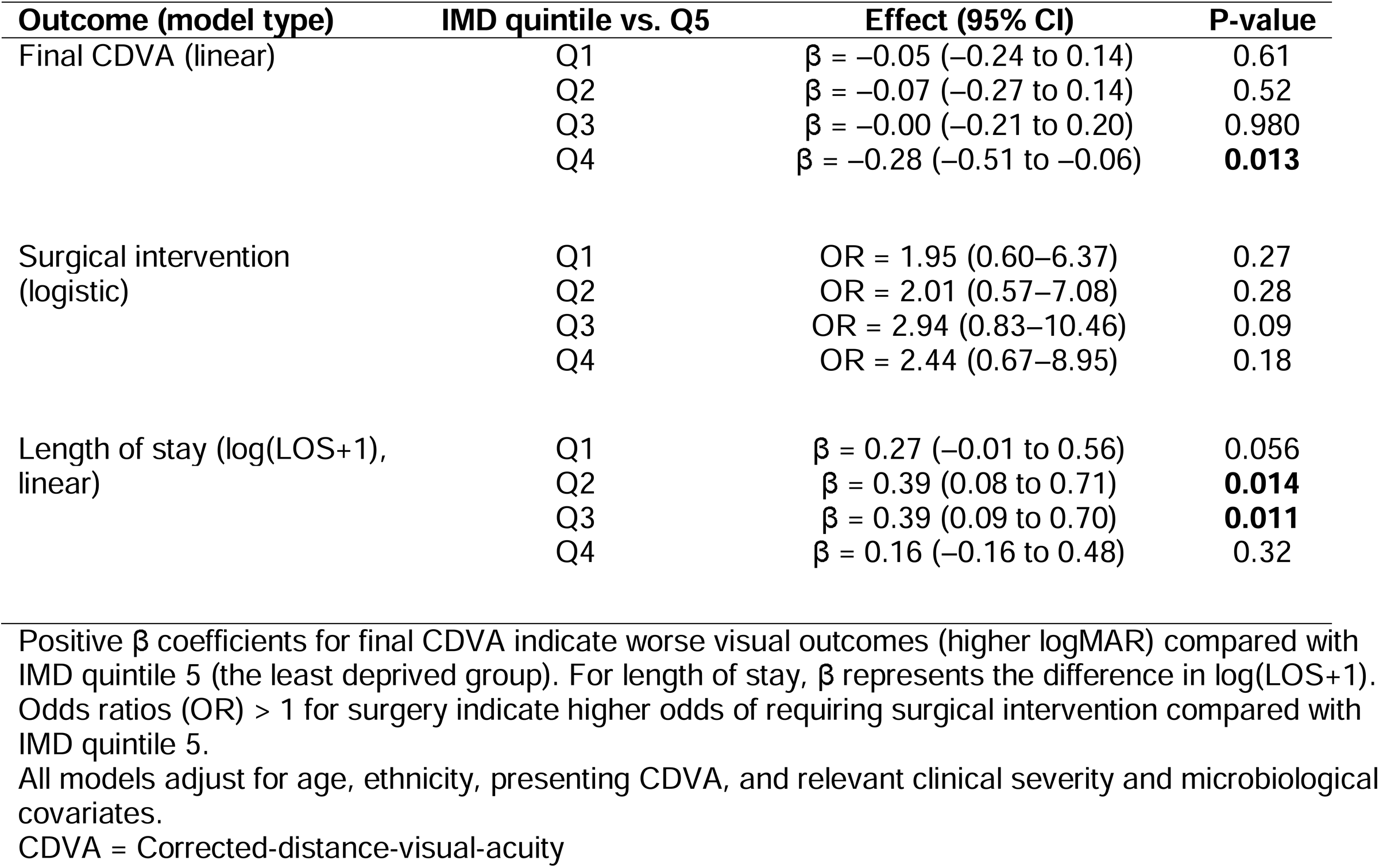
Adjusted associations between Index of Multiple Deprivation (IMD) quintile and clinical outcomes.

Among admitted patients, LOS was modestly longer in IMD Q2 (β=0.392; 95% CI, 0.080–0.705; p=0.014) and IMD Q3 (β=0.393; 95% CI, 0.091–0.695; p=0.011) compared with Q5, although patients in the most deprived quintile (Q1) did not have prolonged stays (β=0.274; 95% CI, – 0.007 to 0.555; p=0.056) (**Supplementary Table S8**). Worse presenting CDVA was associated with longer hospitalisation (β=0.169; 95% CI 0.047–0.292; p=0.007), consistent with the relationship between initial severity and inpatient resource use.

## DISCUSSION

This study provides one of the most comprehensive evaluation of the relationship between socioeconomic deprivation and BK in the UK. Contrary to expectations and trends observed in other ophthalmic diseases, deprivation was not associated with presenting severity, hospital admission, or visual outcomes. Instead, presenting visual acuity, age, and microbiological profile were the main determinants of prognosis. These findings suggest that, within a universally accessible healthcare system, the impact of deprivation on acute ophthalmic emergencies may be reduced, potentially due to the painful and visually alarming nature of BK prompting urgent care-seeking across socioeconomic groups.

Socioeconomic gradients are well-recognised in ophthalmology, with more deprived groups often presenting later and with more severe disease, as reported in glaucoma, diabetic retinopathy, and age-related macular degeneration.^14,15,17,29^ These disparities are typically interpreted through the lens of delayed care-seeking, reduced health literacy, limited access to preventive services, and competing socioeconomic pressures.

In contrast, our study did not demonstrate any deprivation-related differences in presenting CDVA, infiltrate or epithelial defect size, hypopyon, or culture positivity. Although certain predisposing risk factors showed socioeconomic variation such as contact lens wear being more common among less deprived patients and absence of identifiable risk factors more frequent in the most deprived, these differences did not translate into any measurable variation in presenting severity or clinical outcomes. One potential explanation is the acute, painful, and vision-threatening nature of BK, which may prompt urgent care-seeking regardless of socioeconomic status. This is supported by our finding that time to presentation was short (median 3 days, IQR 1–6) and did not differ across IMD groups, suggesting that delays in accessing care were not disproportionately greater among more deprived patients. Rapid-onset symptoms such as pain and visual loss may minimise behavioural variation in help-seeking between social groups. This interpretation is supported by recent UK studies during the COVID-19 pandemic, which demonstrated that despite national lockdowns and major reductions in attendance for other ophthalmic conditions, presentations for microbial keratitis remained stable, likely because pain was the primary driver of attendance.^23,30^ In addition, the presence of open-access emergency eye services and community-optometry triage pathways within the NHS may offer relatively equitable early access across deprivation levels.

Reasons for admission included concerns regarding adherence to intensive hourly topical therapy, limited social support, and ability to safely self-administer treatment. In contrast to national data demonstrating higher unplanned hospitalisation rates among more deprived UK populations across multiple disease areas,^31^ deprivation did not influence hospital admission in this BK cohort. Instead, presenting CDVA was the main determinant, reflecting clinical decision-making based on functional severity. This likely reflects the equalising effect of NHS urgent eye-care pathways, including walk-in services and community-optometry triage. While deprivation may influence BK incidence, management decisions at tertiary level appear driven by clinical severity rather than socioeconomic status. This finding diverges from prior studies suggesting that BK imposes disproportionate economic burden on deprived communities.^22^ It is possible that while socioeconomic disadvantage contributes to higher overall BK incidence at the population level, once patients present to tertiary services, management decisions are driven overwhelmingly by clinical severity rather than deprivation.

Final CDVA similarly showed no association with deprivation. Instead, presenting CDVA, age, and microbiological profile, particularly Gram-positive bacteria, were the strongest predictors of visual prognosis. The relatively high proportion of culture-negative cases (63.7%) included in this study is likely related to the high proportion of cases with mild severity of BK where >60% of the cases had a small epithelial defect or infiltrate. However, the relatively low culture yield is consistent with the findings of the literature, which highlighted high variability of the culture yield depending on the geographical location, study design, patient population, and sampling technique.^32^ The need for surgical intervention was also driven by microbiological and clinical markers of severity, particularly culture positivity with Gram-positive organisms and presence of a large epithelial defect, but not by deprivation. This reinforces that biological rather than socioeconomic factors drive progression to surgical management once patients are under specialist care.

Among admitted patients, LOS showed only minor variation across IMD quintiles, with no significant difference in the most deprived group. Presenting CDVA remained the strongest predictor of LOS. These findings align with previous UK studies demonstrating that inpatient burden in IK is primarily driven by disease severity rather than socioeconomic factors.^5,22^

International evidence has suggested that socioeconomic disadvantage may worsen BK presentation. A recent US analysis reported neighbourhood-level deprivation associated with more severe presenting CDVA, likely reflecting barriers in insurance-based care models.^21^ The absence of such associations in our cohort highlights the potential mitigating effect of the NHS, where emergency ophthalmic care, hospitalisation, and most treatments are provided free at the point of use through the NHS, which may reduce financial barriers to accessing care. This contrasts with insurance-based systems where out-of-pocket costs may influence care-seeking behaviour. Although deprivation has been reported to influence long-term graft survival, ocular trauma outcomes, and chronic ocular disease trajectories,^14,33^ these effects may not extend to acute corneal infections, where urgency overrides socioeconomic variation in presentation timing.

Strengths of this study include its large sample size, multicentre recruitment, linkage with a nationally standardised deprivation index, and comprehensive clinical and microbiological assessment. The use of multivariable models enabled robust adjustment for demographic and biological confounders. Limitations include the retrospective design. IMD is an area-level measure that may mask individual socioeconomic variation. Absence of variables such as medication adherence, and primary-care interactions limits evaluation of behavioural pathways. Small numbers in minority race groups prevented detailed subgroup analyses.

In this UK BK cohort, socioeconomic deprivation was not associated with presenting severity, hospitalisation, or visual outcomes. Instead, clinical severity at presentation, age, and microbiological profile were the main determinants of prognosis. These findings suggest that in BK within a universally accessible healthcare system, the influence of deprivation may be reduced. Further prospective studies incorporating behavioural and access-to-care factors are warranted.

## Data Availability

All data produced in the present study are available upon reasonable request to the authors

## ACKNOWLEDGEMENT

The authors thank all participating ophthalmology departments for data provision and the clinical audit teams for facilitating this service-evaluation study. The authors acknowledge the contributions of Muhammad Nasir (Statistician) in the statistical analysis of the data.

## ETHICS STATEMENT

Ethics approval was not required as this clinical audit only involved retrospective study of anonymised data that were collected during the process of standard clinical care.

## Notes

**Funding/support:** DSJT acknowledges funding support from the Medical Research Council Clinician Scientist Fellowship (UKRI2441) and Birmingham Health Partners (BHP) Clinician Scientist Fellowship.

**Conflict of interest:** None

### Competing Interest Statement

The authors have declared no competing interest.

### Funding Statement

Funding support from the Medical Research Council Clinician Scientist Fellowship (UKRI2441) and Birmingham Health Partners (BHP) Clinician Scientist Fellowship.

### Author Declarations

The study was approved by the clinical governance team of both institutions as a clinical audit (BMEC Ref: 2444; QMC Ref: 19-265C) and was conducted in accordance with the tenets of Declaration of Helsinki. Ethics approval was not required as this clinical audit only involved retrospective study of anonymised data that were collected during the process of standard clinical care.

## REFERENCES

1. Stapleton F, Carnt N. Contact lens-related microbial keratitis: how have epidemiology and genetics helped us with pathogenesis and prophylaxis. Eye (Lond). 2012; 26, 185–93.

2. Cabrera-Aguas M, Khoo P, Watson SL. Infectious keratitis: A review. Clin Exp Ophthalmol. 2022; 50, 543–62.

3. Ting DSJ, Deshmukh R, Ting DSW, Ang M. Big data in corneal diseases and cataract: Current applications and future directions. Front Big Data. 2023; 6, 1017420.

4. Ting DSJ, Cairns J, Gopal BP, Ho CS, Krstic L, Elsahn A, et al. Risk Factors, Clinical Outcomes, and Prognostic Factors of Bacterial Keratitis: The Nottingham Infectious Keratitis Study. Front Med (Lausanne). 2021; 8, 715118.

5. Hossain P. Microbial keratitis-the true costs of a silent pandemic? Eye (Lond). 2021; 35, 2071–2.

6. Ibrahim YW, Boase DL, Cree IA. Epidemiological characteristics, predisposing factors and microbiological profiles of infectious corneal ulcers: the Portsmouth corneal ulcer study. Br J Ophthalmol. 2009; 93, 1319–24.

7. Suresh L, Hammoudeh Y, Ho CS, Ong ZZ, Cairns J, Gopal BP, et al. Clinical features, risk factors and outcomes of contact lens-related bacterial keratitis in Nottingham, UK: a 7-year study. Eye (Lond). 2024; 38, 3459–66.

8. Ting DSJ, Ho CS, Deshmukh R, Said DG, Dua HS. Infectious keratitis: an update on epidemiology, causative microorganisms, risk factors, and antimicrobial resistance. Eye (Lond). 2021; 35, 1084–101.

9. Marmot M. Social determinants of health inequalities. Lancet. 2005; 365, 1099–104.

10. Bambra C. Health inequalities and welfare state regimes: theoretical insights on a public health ‘puzzle’. J Epidemiol Community Health. 2011; 65, 740–5.

11. England L, O’Connor A. Do Socioeconomic Inequalities Exist Within Ophthalmology and Orthoptics in the UK?: A Scoping Review. Br Ir Orthopt J. 2024; 20, 31–47.

12. Green MA, Daras K, Davies A, Barr B, Singleton A. Developing an openly accessible multi-dimensional small area index of ‘Access to Healthy Assets and Hazards’ for Great Britain, 2016. Health & Place. 2018; 54, 11–9.

13. Barlow P, Mohan G, Nolan A, Lyons S. Area-level deprivation and geographic factors influencing utilisation of General Practitioner services. SSM Popul Health. 2021; 15, 100870.

14. Lane M, Lane V, Abbott J, Braithwaite T, Shah P, Denniston AK. Multiple deprivation, vision loss, and ophthalmic disease in adults: global perspectives. Surv Ophthalmol. 2018; 63, 406–36.

15. Wong TL, Ang JL, Deol S, Buckmaster F, McTrusty AD, Tatham AJ. The relationship between multiple deprivation and severity of glaucoma at diagnosis. Eye (Lond). 2023; 37, 3376–81.

16. Ng WS, Agarwal PK, Sidiki S, McKay L, Townend J, Azuara-Blanco A. The effect of socio-economic deprivation on severity of glaucoma at presentation. Br J Ophthalmol. 2010; 94, 85–7.

17. Low L, Law JP, Hodson J, McAlpine R, O’Colmain U, MacEwen C. Impact of socioeconomic deprivation on the development of diabetic retinopathy: a population-based, cross-sectional and longitudinal study over 12 years. BMJ Open. 2015; 5, e007290.

18. Acharya N, Lois N, Townend J, Zaher S, Gallagher M, Gavin M. Socio-economic deprivation and visual acuity at presentation in exudative age-related macular degeneration. Br J Ophthalmol. 2009; 93, 627–9.

19. Ting DSJ, Galal M, Kulkarni B, Elalfy MS, Lake D, Hamada S, et al. Clinical Characteristics and Outcomes of Fungal Keratitis in the United Kingdom 2011-2020: A 10-Year Study. J Fungi (Basel). 2021; 7.

20. Prajna NV, Krishnan T, Rajaraman R, Patel S, Shah R, Srinivasan M, et al. Predictors of Corneal Perforation or Need for Therapeutic Keratoplasty in Severe Fungal Keratitis: A Secondary Analysis of the Mycotic Ulcer Treatment Trial II. JAMA Ophthalmol. 2017; 135, 987–91.

21. Hicks PM, Niziol LM, Newman-Casey PA, Salami K, Singh K, Woodward MA. Social Risk Factor Associations With Presenting Visual Acuity in Patients With Microbial Keratitis. JAMA Ophthalmol. 2023; 141, 727–34.

22. Moussa G, Hodson J, Gooch N, Virdee J, Penaloza C, Kigozi J, et al. Calculating the economic burden of presumed microbial keratitis admissions at a tertiary referral centre in the UK. Eye (Lond). 2021; 35, 2146–54.

23. Butt GF, Recchioni A, Moussa G, Hodson J, Wallace GR, Murray PI, et al. The impact of the COVID-19 pandemic on microbial keratitis presentation patterns. PLoS One. 2021; 16, e0256240.

24. Ministry of Housing C, Local G. The English Indices of Deprivation 2019 (IoD2019). London: HM Government; 2019 2019.

25. Silvester A, Scott R, Pitalia A. Impact of social deprivation on cataract presentation and surgical outcome in the north of England: a retrospective cohort study. The Lancet. 2018; 392, S81.

26. Knight A, Lindfield R. The relationship between socio-economic status and access to eye health services in the UK: a systematic review. Public Health. 2015; 129, 94–102.

27. Moussa G, Kalogeropoulos D, Ch’ng SW, Lett KS, Mitra A, Tyagi AK, et al. Effect of deprivation and ethnicity on primary macula-on retinal detachment repair success rate and clinical outcomes: A study of 568 patients. PLoS One. 2021; 16, e0259714.

28. Ting DSJ, Ho CS, Cairns J, Gopal BP, Elsahn A, Al-Aqaba M, et al. Seasonal patterns of incidence, demographic factors and microbiological profiles of infectious keratitis: the Nottingham Infectious Keratitis Study. Eye (Lond). 2021; 35, 2543–9.

29. Yip JL, Luben R, Hayat S, Khawaja AP, Broadway DC, Wareham N, et al. Area deprivation, individual socioeconomic status and low vision in the EPIC-Norfolk Eye Study. J Epidemiol Community Health. 2014; 68, 204–10.

30. De Sousa Peixoto R, Lakhani BK, Xue Y, Dua HS, Said DG, King AJ. Pain, the driving force behind eye casualty attendance during the COVID-19 lockdown. Indian J Ophthalmol. 2020; 68, 2309–10.

31. Agency UKHS. Health Inequalities in Health Protection Report 2025. London, UK: UK Government; 2025 2025.

32. Ting DSJ, Gopal BP, Deshmukh R, Seitzman GD, Said DG, Dua HS. Diagnostic armamentarium of infectious keratitis: A comprehensive review. Ocul Surf. 2022; 23, 27–39.

33. Chua PY, Azuara-Blanco A, Hulme W, Jones MNA, Mustafa MS, Kaye SB. The effect of socioeconomic deprivation on corneal graft survival in the United Kingdom. Ophthalmology. 2013; 120, 2436–41.

