## Supplementary Tables for "Impact of Socioeconomic Status on Clinical Features and Outcomes of Bacterial Keratitis: The Midlands Infectious Keratitis Study"

### Supplementary Table S1. Full multivariable linear regression model evaluating factors associated with presenting corrected-distance-visual-acuity (CDVA) in logMAR unit.

| **Parameters** | **Estimate (β)** | **95% CI lower** | **95% CI upper** | **p-value** |
| --- | --- | --- | --- | --- |
| Intercept | 0.129 | -0.296 | 0.555 | 0.55 |
| Age (per year) | 0.017 | 0.011 | 0.023 | **<0.001** |
| Sex (Reference: Male) | | | | |
| Female | -0.030 | -0.268 | 0.207 | 0.80 |
| IMD quintile (Reference: Q5) | | | | |
| IMD Q1 vs Q5 | 0.243 | -0.095 | 0.580 | 0.16 |
| IMD Q2 vs Q5 | 0.038 | -0.321 | 0.396 | 0.84 |
| IMD Q3 vs Q5 | 0.132 | -0.226 | 0.491 | 0.47 |
| IMD Q4 vs Q5 | 0.096 | -0.289 | 0.480 | 0.63 |

### CDVA = Corrected-distance-visual-acuity; IMD = Index of Multiple Deprivation.

### Supplementary Table S2. Full multivariable logistic regression model evaluating predictors of a large corneal infiltrate at presentation.

| **Parameters** | **OR** | **95% CI lower** | **95% CI upper** | **p-value** |
| --- | --- | --- | --- | --- |
| Intercept | 0.003 | 0.000 | 0.025 | <0.001 |
| Age (per year) | 1.002 | 0.981 | 1.024 | 0.83 |
| IMD quintile (Reference: Q5) | | | | |
| IMD Q1 vs Q5 | 2.081 | 0.593 | 7.304 | 0.25 |
| IMD Q2 vs Q5 | 0.932 | 0.215 | 4.042 | 0.93 |
| IMD Q3 vs Q5 | 1.520 | 0.380 | 6.081 | 0.55 |
| IMD Q4 vs Q5 | 2.844 | 0.700 | 11.562 | 0.14 |
| Presenting CDVA | 6.095 | 3.282 | 11.319 | <0.001* |

### OD = Odds ratio; CDVA = Corrected-distance-visual-acuity; IMD = Index of Multiple Deprivation

### Supplementary Table S3. Full multivariable logistic regression model investigating predictors of a large epithelial defect at presentation.

| **Parameters** | **OR** | **95% CI lower** | **95% CI upper** | **p-value** |
| --- | --- | --- | --- | --- |
| Intercept | 0.007 | 0.001 | 0.043 | **<0.001** |
| Age (per year) | 0.996 | 0.977 | 1.015 | 0.69 |
| IMD quintile (Reference: Q5) | | | | |
| IMD Q1 vs Q5 | 2.031 | 0.594 | 6.944 | 0.26 |
| IMD Q2 vs Q5 | 2.510 | 0.680 | 9.266 | 0.17 |
| IMD Q3 vs Q5 | 2.595 | 0.704 | 9.562 | 0.15 |
| IMD Q4 vs Q5 | 3.273 | 0.846 | 12.666 | 0.09 |
| Presenting CDVA | 5.000 | 3.069 | 8.147 | **<0.001** |

### OR = Odds ratio; CDVA = Corrected-distance-visual-acuity; IMD = Index of Multiple Deprivation

**Supplementary Table S4.** Full multivariable logistic regression model assessing factors associated with the presence of hypopyon at presentation.

| **Parameters** | **OR** | **95% CI lower** | **95% CI upper** | **p-value** |
| --- | --- | --- | --- | --- |
| Intercept | 0.098 | 0.036 | 0.268 | **<0.001** |
| Age (per year) | 1.012 | 0.998 | 1.026 | 0.09 |
| IMD quintile (Reference: Q5) | | | | |
| IMD Q1 vs Q5 | 1.142 | 0.523 | 2.491 | 0.74 |
| IMD Q2 vs Q5 | 0.829 | 0.355 | 1.938 | 0.67 |
| IMD Q3 vs Q5 | 0.733 | 0.306 | 1.755 | 0.49 |
| IMD Q4 vs Q5 | 1.118 | 0.450 | 2.778 | 0.81 |
| Presenting CDVA | 1.798 | 1.368 | 2.364 | **<0.001** |

OR = Odds ratio; CDVA = Corrected**-**distance**-**visual**-**acuity; IMD = Index of Multiple Deprivation

### Supplementary Table S5. Full multivariable logistic regression model identifying factors associated with hospitalization for bacterial keratitis.

| **Parameters** | **OR** | **95% CI lower** | **95% CI upper** | **P-value** |
| --- | --- | --- | --- | --- |
| Intercept | 0.420 | 0.180 | 0.979 | 0.04 |
| Age (per year) | 1.008 | 0.996 | 1.021 | 0.18 |
| IMD quintile (Reference: Q5) | | | | |
| IMD Q1 vs Q5 | 1.141 | 0.562 | 2.314 | 0.72 |
| IMD Q2 vs Q5 | 0.693 | 0.327 | 1.469 | 0.34 |
| IMD Q3 vs Q5 | 0.882 | 0.414 | 1.876 | 0.74 |
| IMD Q4 vs Q5 | 1.205 | 0.525 | 2.769 | 0.66 |
| Presenting CDVA | 1.693 | 1.309 | 2.188 | **<0.001** |

### OR = Odds ratio; CDVA = Corrected-distance-visual-acuity; IMD = Index of Multiple Deprivation

**Supplementary Table S6.** Complete multivariable linear regression model examining predictors of final corrected-distance-visual-acuity (CDVA) in logMAR unit.

| **Parameters** | **Estimate (β)** | **95% CI lower** | **95% CI upper** | **p-value** |
| --- | --- | --- | --- | --- |
| Intercept | -0.567 | -1.051 | -0.083 | 0.02 |
| Age (per year) | 0.008 | 0.005 | 0.012 | **<0.001** |
| IMD quintile (Reference: Q5) | | | | |
| IMD Q1 vs Q5 | -0.050 | -0.242 | 0.143 | 0.61 |
| IMD Q2 vs Q5 | -0.067 | -0.272 | 0.139 | 0.52 |
| IMD Q3 vs Q5 | -0.003 | -0.208 | 0.203 | 0.98 |
| IMD Q4 vs Q5 | -0.284 | -0.507 | -0.061 | **0.013** |
| Presenting CDVA | 0.628 | 0.545 | 0.710 | **<0.001** |
| Infiltrate size (Reference: Small) | | | | |
| Large | 0.285 | -0.084 | 0.653 | 0.13 |
| Moderate | 0.118 | -0.135 | 0.371 | 0.36 |
| ED size (Reference: Small) | | | | |
| Large | 0.112 | -0.234 | 0.458 | 0.53 |
| Moderate | 0.023 | -0.228 | 0.274 | 0.86 |
| Hypopyon (Reference: No) | | | | |
| Yes | -0.059 | -0.219 | 0.101 | 0.47 |
| Culture (Reference: Negative) | | | | |
| Positive | -0.005 | -0.399 | 0.389 | 0.98 |
| Organism (Reference: Gram-negative) | | | | |
| Gram-positive | 0.299 | 0.076 | 0.521 | **0.009** |

CDVA = Corrected**-**distance**-**visual**-**acuity; IMD = Index of Multiple Deprivation; ED = Epithelial defect.

### Supplementary Table S7. Full multivariable logistic regression model assessing predictors of requiring surgical intervention.

| **Parameters** | **OR** | **95% CI lower** | **95% CI upper** | **p-value** |
| --- | --- | --- | --- | --- |
| Intercept | 0.000 | 0.000 | 0.005 | <0.001 |
| Age (per year) | 1.019 | 0.999 | 1.040 | 0.06 |
| IMD quintile (Reference: Q5) | | | | |
| IMD Q1 vs Q5 | 1.947 | 0.595 | 6.370 | 0.27 |
| IMD Q2 vs Q5 | 2.014 | 0.573 | 7.078 | 0.28 |
| IMD Q3 vs Q5 | 2.941 | 0.827 | 10.455 | 0.09 |
| IMD Q4 vs Q5 | 2.442 | 0.667 | 8.945 | 0.18 |
| Presenting CDVA | 1.830 | 1.174 | 2.853 | **0.008** |
| Infiltrate size (Reference: small) | | | | |
| Large | 1.422 | 0.307 | 6.591 | 0.65 |
| Moderate | 1.022 | 0.300 | 3.479 | 0.97 |
| ED size (Reference: small) | | | | |
| Large | 5.288 | 1.187 | 23.565 | **0.029** |
| Moderate | 3.323 | 0.912 | 12.109 | 0.07 |
| Hypopyon (Reference: No) | | | | |
| Yes | 1.111 | 0.490 | 2.519 | 0.80 |
| Culture (Reference: Negative) | | | | |
| Positive | 13.999 | 1.013 | 193.507 | **0.049** |
| Organism (Reference: Gram-negative) | | | | |
| Gram-positive | 3.982 | 1.193 | 13.294 | **0.025** |

### OR = Odds ratios; CDVA = corrected distance visual acuity; IMD = Index of Multiple Deprivation; ED = epithelial defect.

**Supplementary Table S8.** Complete multivariable linear regression model evaluating predictors of duration of hospitalization, analyzed as log(LOS + 1).

| **Parameters** | **Estimate (β)** | **95% CI lower** | **95% CI upper** | **p-value** |
| --- | --- | --- | --- | --- |
| Intercept | 1.274 | 0.353 | 2.195 | 0.01 |
| Age (per year) | 0.005 | -0.000 | 0.010 | 0.051 |
| IMD quintile (Reference: Q5) | | | | |
| IMD Q1 vs Q5 | 0.274 | -0.007 | 0.555 | 0.06 |
| IMD Q2 vs Q5 | 0.392 | 0.080 | 0.705 | **0.014** |
| IMD Q3 vs Q5 | 0.393 | 0.091 | 0.695 | **0.011** |
| IMD Q4 vs Q5 | 0.163 | -0.159 | 0.484 | 0.32 |
| Presenting CDVA | 0.169 | 0.047 | 0.292 | **0.007** |
| Infiltrate size (Reference: Small) | | | | |
| Large | 0.591 | -0.016 | 1.199 | 0.06 |
| Moderate | 0.335 | -0.033 | 0.703 | 0.07 |
| ED size (Reference: Small) | | | | |
| Large | -0.377 | -0.950 | 0.196 | 0.19 |
| Moderate | -0.149 | -0.518 | 0.221 | 0.43 |
| Hypopyon (Reference: No) | | | | |
| Yes | 0.092 | -0.126 | 0.310 | 0.41 |
| Culture (Reference: Negative) | | | | |
| Positive | -0.271 | -1.037 | 0.495 | 0.49 |
| Organism (Reference: Gram-negative) | | | | |
| Gram-positive | 0.213 | -0.062 | 0.489 | 0.13 |

CDVA = corrected distance visual acuity; IMD = Index of Multiple Deprivation; ED = epithelial defect.
